# Antibody titers against the Alpha, Beta, Gamma, and Delta variants of SARS-CoV-2 induced by BNT162b2 vaccination measured using automated chemiluminescent enzyme immunoassay

**DOI:** 10.1101/2021.09.23.21263927

**Authors:** Hideaki Kato, Kei Miyakawa, Norihisa Ohtake, Hirofumi Go, Yutaro Yamaoka, Satoshi Yajima, Tomoko Shimada, Atsushi Goto, Hideaki Nakajima, Akihide Ryo

## Abstract

**Background:** Levels of 50% neutralizing titer (NT50) reflect a vaccine-induced humoral immunity after the vaccination against the severe acute respiratory syndrome coronavirus-2 (SARS-CoV-2). Measurements of NT50 are difficult to implement in large quantities. A high-throughput laboratory test is expected for determining the level of herd immunity against SARS-CoV-2.

**Methods:** We analyzed samples from 168 Japanese healthcare workers who had completed two doses of the BNT162b2 vaccine. We analyzed immunoglobulin G (IgG) index values against spike protein (SP) using automated chemiluminescent enzyme immunoassay system AIA-CL and analyzed the background factors affecting antibody titer. SP IgG index was compared with 50% neutralization titers.

**Results:** The median SP IgG index values of the subjects (mean age = 43 years; 75% female) were 0.1, 1.35, 60.80, and 97.35 before and at 2, 4, and 6 weeks after the first dose, respectively. At 4 and 6 weeks after the first dose, SP IgG titers were found to have positive correlation with NT50 titer (r=0.7535 in 4 weeks; r=0.4376 in 6 weeks). Proportions of the SP IgG index values against the Alpha, Beta, Gamma, and Delta variants compared with the original strain were 2.029, 0.544, 1.017, and 0.6096 respectively. Older age was associated with lower SP IgG titer index 6 weeks after the first dose.

**Conclusions:** SP IgG index values were raised at 3 weeks after two doses of BNT162b2 vaccination and have positive correlation with NT50. SP IgG index values were lower in the older individuals and against Beta and Delta strain.

## Introduction

The coronavirus disease 2019 (COVID-19) caused by the severe acute respiratory syndrome coronavirus 2 (SARS-CoV-2) is a global public health threat that has infected 209 million people and caused 4.3 million deaths as of August 19, 2021. The messenger RNA vaccine gives rapid and robust immunity comparable to natural infection by the virus [1]. The vaccine, which is the cornerstone of current control strategies, was designed to primarily target the SARS-CoV-2 spike protein (SP) of the prototype Wuhan strain [2]. The mRNA vaccines (BNT162b22 and mRNA-1273) elicit high titers of SARS-CoV-2 neutralizing antibodies [3]. Consequently, the efficacy of these vaccines in preventing illness and reducing disease severity range from 94% to 95% [4,5]. Determining the level of herd immunity against COVID-19 requires mass surveillance of the immune response to SARS-CoV-2 and its variants among vaccinated individuals. Khoury et al. showed that the titers of vaccine-induced neutralizing antibody responses are highly predictive of immune protection [6]. Moreover, measuring the levels of neutralizing antibodies helps to determine the status of an individual’s protective immunity against virus infection. Viral neutralization titers measure the ability of antibodies to prevent viral infection of a eukaryotic cell line in vitro. Although the current gold standard neutralization assay requires live SARS-CoV-2, pseudotyped virus-based assays have also been commonly used because of their safety and versatility [7]. However, these conventional neutralization assays are of relatively low throughput and require long readout times. Overcoming these limitations requires the development of alternative means for measuring surrogate antibodies that can replace the cell-mediated neutralization assay. Recently, we developed a dedicated reagents against SARS-CoV-2 spike protein using an automated chemiluminescent enzyme immunoassay (CLEIA) system AIA-CL (TOSOH, Japan). It is a high-throughput serological test capable of processing 120 samples per hour. It simultaneously detects immunoglobulin G (IgG) against the nucleocapsid protein (NP) and SP of SARS-CoV-2 [8]. In tests on COVID-19 patients, an automated chemiluminescent enzyme immunoassay system using the AIA-CL system exhibited 100% sensitivity and specificity in detecting antibodies against SARS-CoV-2. Here, we sought to elucidate the relationship between the level of binding antibodies measured by the CLEIA and neutralization activity determined by a pseudovirus-based neutralization assay in a cohort of donors without a past history of COVID-19. Our aims were, first, to determine the usefulness the commercial CLEIA reagents using the AIA-CL system in assessing vaccine-induced changes in SP-specific IgG (SP IgG) levels against SARS-CoV-2 after BNT162b2 vaccination and comparing these results with the corresponding 50% neutralization titers (NT50); second, to analyze the SP IgG index titers between the original strain and variants of SARS-CoV-2 after BNT162b2 mRNA vaccination using the CLEIA using AIA-CL system designed for the original and these variants; and third, to identify the underlying host factors determining vaccine-elicited humoral immunity, as these factors remain unclear in Asian populations.

### Patients and method

Participants were recruited in March 2021 from among the 1800 healthcare workers in the Yokohama City University Hospital. The hospital has 672 admission beds, including 21 beds for COVID-19 patients. Blood samples were collected from the following four time points: prior to vaccination and then at 2, 4, and 6 weeks after the first dose (the second dose was administered 3 weeks after the first dose). The actual collection day ranged within 3 days before and after the scheduled collection day. The first vaccine dose was administered between March 15 and 22, and the second dose was administered between April 5 and 13. Subjects were asked to provide the following information: birthday and year, sex, drinking habit, smoking habit, comorbidities, body height, and body weight. The participants’ age and body mass index at the first dose was calculated. Alcohol drinking habit was categorized corresponding to never drink; occasionally drink or drink several times a week; and drink daily. Smoking habit was categorized corresponding to never smoke, ex-smoker, and current smoker. Comorbidities were defined as having chronic respiratory diseases (including bronchial asthma, diabetes mellitus) and currently undergoing antitumor chemotherapy or immunosuppression, regardless of the doses and types of regimens.

### Chemiluminescent enzyme immunoassay for SARS-CoV-2 SP RBD-IgG on AIA-CL1200

The index value of SARS-CoV-2 SP and NP IgG were measured using the commercial CLEIA reagents (AIA-CL SARS-CoV-2 SP-IgG antibody detection reagent, Tosoh, Japan). Previous validations of these reagents have shown this setting to have high sensitivity and specificity [8]. Furthermore, to compare the IgG index values between the original and variant RBD, the original strain, the Alpha (N501Y, B.1.1.7), Beta (K417N, E484K and N501Y, B.1.351), Gamma (K417T, E484K and N501Y, P.1), and Delta (D614G, T478K, P681R and L452R) RBD Flag-tagged recombinant proteins were expressed in Expi293F cells (Thermo Fisher Scientific, Waltham, USA), according to the manufacturer’s instructions. The supernatants containing the recombinant proteins were collected and purified using the Strep-tag purification system (IBA Lifesciences). Purified these recombinant RBD protein from each variant was immobilized to micromagnetic beads respectively and prepared the prototype reagent panel for each strain on AIA-CL system and their attachment to the AIA-CL instrument have been described previously. The intensities of the chemiluminescent signals from 100 healthy donor samples were measured with cutoff values (1.0 index value) determined from the mean + 6SD.

### Neutralizing assay using a pseudovirus

The neutralizing assay using an HIV-based pseudovirus bearing the SARS-CoV-2 spike was performed as described previously [9]. Briefly, VeroE6/TMPRSS2 cells were inoculated with pseudoviruses encoding the luciferase reporter gene within mixtures containing five-fold serially diluted serum (1:50 to 1:31250 dilution). Forty-eight hours after inoculation, the luciferase activity of VeroE6/TMPRSS2 cells was measured using the GloMax Discover System (Promega). We calculated 50% neutralizing titer (NT50) values using Image J software (NIH). In cases where the serum had no observable neutralizing activity to interpolate NT50, we assigned an NT50 value of 25. All samples were assayed in at least duplicate.

### Data analysis

Continuous data are presented either as means with 95% confidence intervals (CIs) or medians with interquartile ranges. Categorical data are presented as numbers and percentages. Continuous variables between two groups were compared using the two-tailed Mann–Whitney U-test, and the Kruskal–Wallis test was used to compare three or more groups. Categorical data were compared using Fisher’s exact test. SP IgG and NT50 antibody titers were correlated using Spearman’s correlation analysis at 4 and 6 weeks after the first dose. A multivariable regression model was used to investigate the association between the variables and SP IgG index values. SP IgG index values were log-transformed for analysis to remove positive skewness. Statistical analyses were performed using Prism 9.1 (GraphPad Software, San Diego, CA, USA) and JMP Pro 15 software (SAS Institute, Cary, NC, USA). *P* values of <0.05 were considered statistically significant. Each of the subjects provided written informed consent before participating in this study. This study was approved by the institutional review board of Yokohama City University Hospital (Approval number: B210300001).

## Results

Written informed consent was obtained from the 189 subjects to participate this study. Among them, 168 subjects had completed blood drawn at all time points. Table 1 shows the basic characteristics of the 168 subjects. Subjects’ median age in years was 43 [35–49] (range 23–62 years). The subjects included 42 males and 126 females. The subjects’ comorbidities were reported as follows: none = 146, bronchial asthma = 16, diabetes mellitus = 2, undergoing antitumor chemotherapy or immunosuppressive therapy = 1, and with chronic lung disease = 1. All participants were Japanese. None of the participants reported having COVID-19, which was confirmed by negative test results for SARS-CoV-2 NP IgG. A plot of SP IgG index values through time is shown in Fig. 1. Before and then at 2, 4, and 6 weeks after the first dose, the SP IgG index values were 0.1 [0.1–0.1] (range 0.0 to 0.4), 1.35 [0.6– 3.25] (range 0.1 to 45.1), 60.80 [26.18–102.8] (range 0.2 to 449.6), and 97.35 [51.43–167.4] (range 3.8 to 927.6), respectively. All subjects exhibited SP IgG index values below the cutoff value of 1.0 before the vaccination.

**Table 1.** Basic characteristics of the subjects, and their SP IgG antibody titers at 3 weeks after the 6 weeks after the first dose. SP IgG index titers were measured using the automated chemiluminescent enzyme immunoassays using AIA-CL system.

|  |  |  | SP IgG index titer (Mean [IQR], range) | *P* values † |
| --- | --- | --- | --- | --- |
| Age | Year old (median [IQR] (range) | 43 [35, 49] (23–62) |  |  |
| Gender |  |  |  |  |
|  | Male, N (%) | 42 (25.0) | 81.4 [45.8, 150.6] (15.0–377.9) | 0.1952 |
|  | Female, N (%) | 126 (75.0) | 97.7 [53.4, 173.2] (3.8–927.6) |  |
| Drinking habit |  |  |  |  |
|  | None, N (%) | 53 (31.5) | 111.9 [54.1, 164.8] (3.8–793.2) | 0.0723 |
|  | By chance, N (%) | 88 (52.4) | 97.9 [51.8, 200.9] (51.8–927.6) |  |
|  | Everyday, N (%) | 27 (16.1) | 79.9 [44.1, 186] (25.5–411.5) |  |
| Smoking habit |  |  |  |  |
|  | None, N (%) | 132 (78.6) | 97.35 [51.08, 163.7] (3.8–927.6) | 0.4442 |
|  | Ex-smoker, N (%) | 29 (17.3) | 111 [56.8, 231.1] (10.8–682.7) |  |
|  | Current smoker, N (%) | 7 (4.2) | 51.8 [45.6, 140] (40.6–222.6) |  |
| Having comorbidities |  |  |  |  |
|  | None, N (%) | 146 (86.9) | 97.35 [51.23, 170.3] (3.8–927.6) | 0.5935 |
|  | Any comorbidities, N (%) | 22 (13.1) | 107.8 [50.93, 151] (36.1–682.7) |  |
| Body mass index | (median [IQR], range) | 21.6 [19.8, 24.2] (16.6–35.6) |  |  |

† SP IgG index titers in the groups were compared using the Mann-Whitney U-test or Kruskal-Wallis test.

**Fig. 1.**
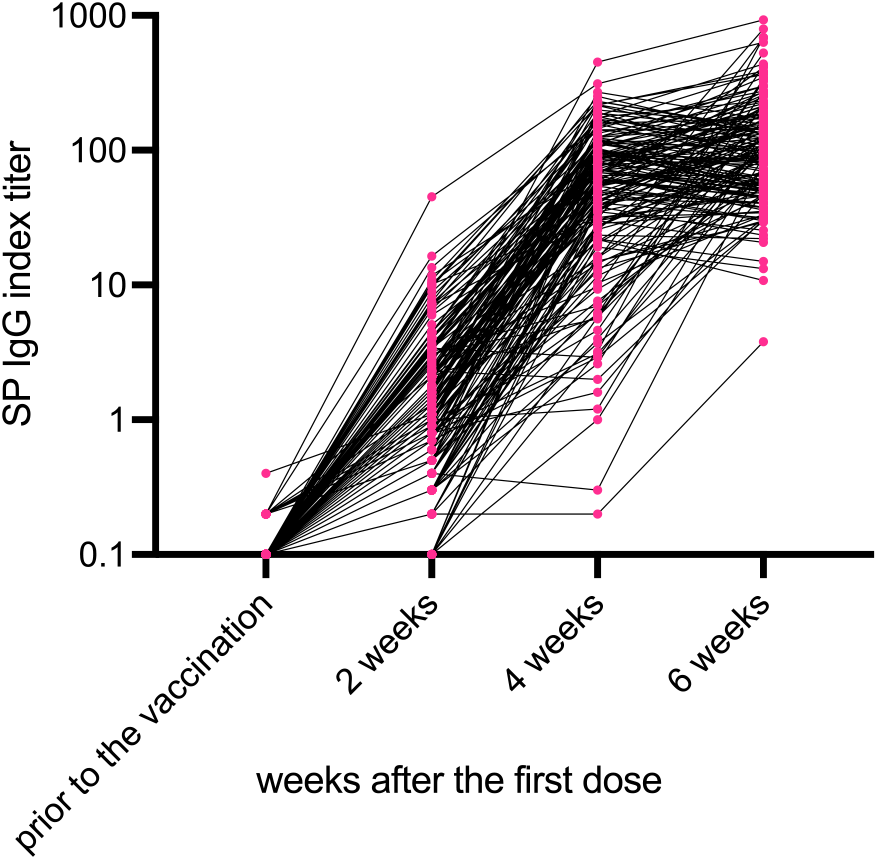
SP IgG titers measured by the AIA-CL system before and then at 2, 4, and 6 weeks after the first dose in the subjects who underwent two doses of BNT162b2 vaccine. Before vaccination, SP IgG titers of all subjects were below the threshold (index 1.0). SP IgG index titers had increased in 2, 4 and 6 weeks after the first dose. Abbreviations: SP, spike protein; IgG, immunoglobulin.

The correlations between NT50 neutralizing antibody titers and AIA-CL SP IgG index values at 4 and 6 weeks after the first dose are shown in Fig. 2 (A) and (B). The NT50 values of neutralizing activity at 4 and 6 weeks after the first vaccine dose are 783.6 [351.2–1389] (range 69.57 to 10073) and 629.8 [386.8–1148] (range 162.6 to 8933), respectively. The NT50 value after two doses of the BNT162b2 vaccine is known to reach more than 100 [10,11]. In our study cohort, 167 subjects (99.4%) had NT50 titers equivalent to or higher than 100 at 4 weeks after the first dose, and all subjects reached this threshold at 6 weeks after the first dose. In terms of SP IgG index values, 151 and 167 subjects (89.9% and 99.4%) had SP IgG index of ≥ 10 at 4 and 6 weeks after the first dose, respectively. SP IgG index values were moderately correlated with NT50 values (Spearman’s correlation coefficient, r = 0.7535 and 0.4376 at 4 and 6 weeks after the first dose, respectively)

**Fig. 2.**
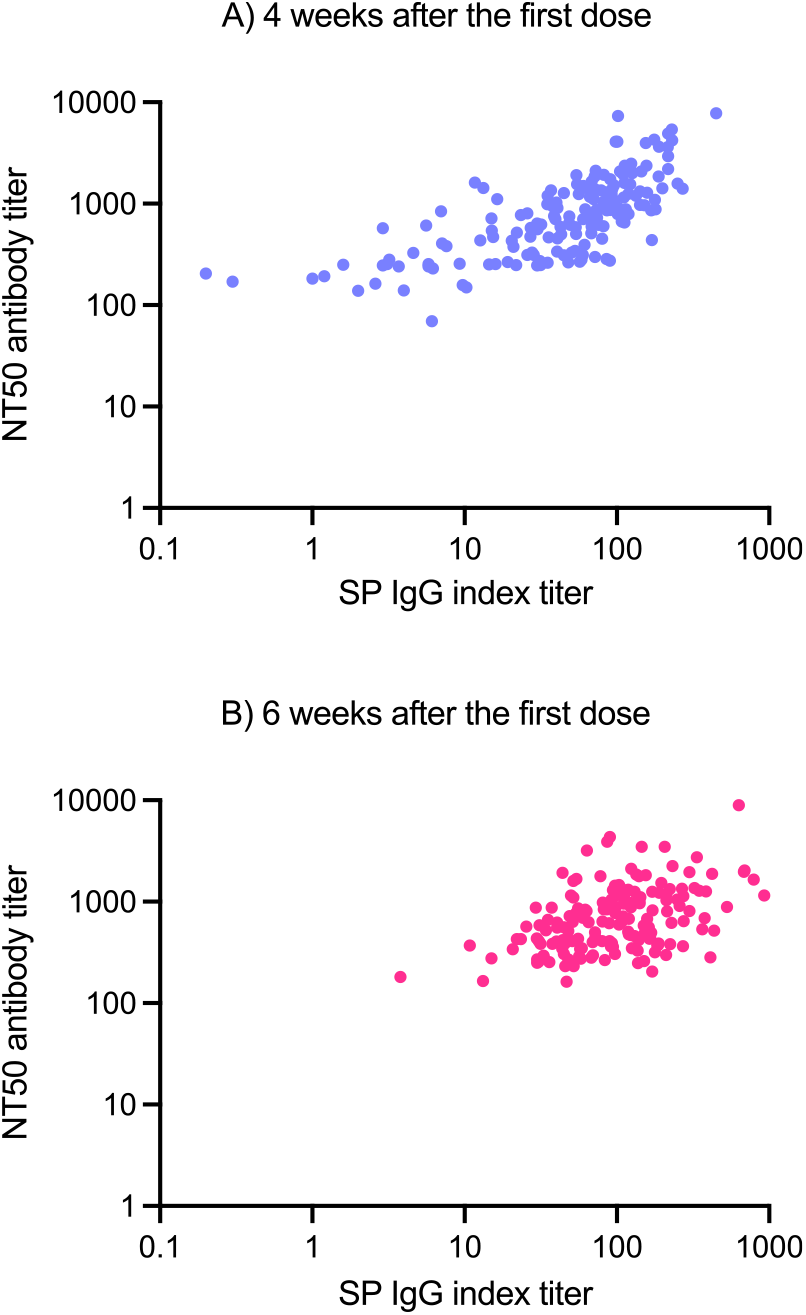
Correlation between NT50 neutralizing antibody titers and AIA-CL SP IgG titer at 4 (A) and 6 (B) weeks after the first dose in the subjects who underwent two doses of BNT162b2 vaccine.

### Comparison of SP IgG index values against the original and the variant strains after vaccination

We compared SP IgG index values against the original (Wuhan) and three variant strains after vaccination. Our results show that the SP IgG index values binding to the Gamma strain (P.1) were similar to or higher than those of the original strain (Fig. 3). However, SP IgG index values against the Beta (B.1.351) and Delta (B.1.617.2) strain were significantly lower than the original strain in sera at 6 weeks after the first dose of the BNT162b2 vaccine. The SP IgG index values against the Alpha (B.1.1.7) strain were significantly higher than that of the original strain at the 6 weeks after the first dose. As the proportions compared to the original strain, the indexes against the Alpha, Beta, Gamma, and Delta strains were 2.029 [1.736–2.359] (*P* < 0.001), 0.544 [0.425–0.660] (*P* < 0.001), 1.017 [0.803–1.220] (*P* < 0.001), and 0.6096 [0.5428–0.7010] (*P* < 0.001) respectively.

**Fig. 3.**
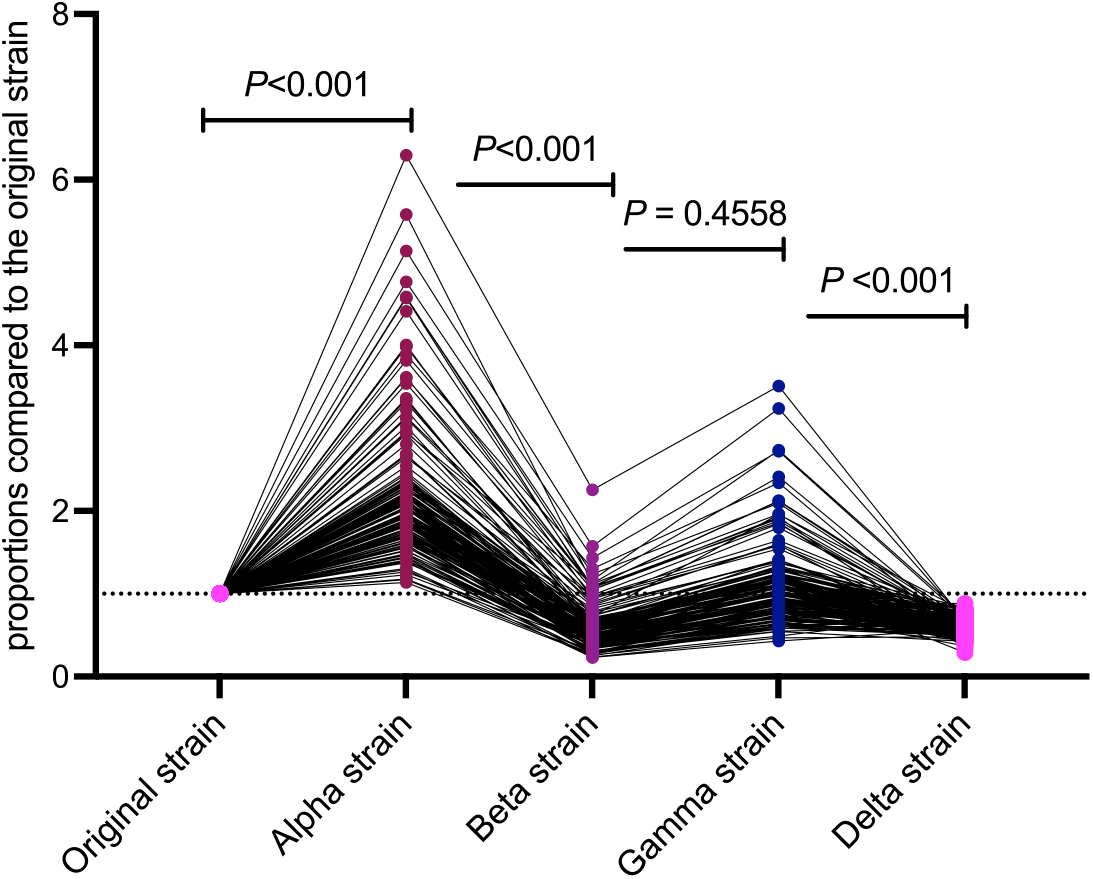
Proportions of antibody titers against the original, Alpha, Beta, Gamma, and Delta strains at 6 weeks after the first dose.

### Background characteristics affecting SP IgG index titer using AIA-CL in the fully vaccinated subjects

With regard to background conditions, there was no significant difference between the SP IgG index values in the subjects according to sex, drinking habit, smoking habit and having any comorbidities (Table 1). The relationship between SP IgG index values and age and body mass index were shown in Fig. 4. We conducted a multivariable regression analysis selecting as background conditions age (year old), sex, drinking habit, smoking habit, having any comorbidities and body mass index. Our model showed that older age affected the log SP IgG index titer in the subjects at 6 weeks after the first vaccine dose (−0.015 [95% CI −0.029, −0.001] in 1-year increments, *P* =0.034). The coefficients of the other covariates are as follows: female gender compared with male (0.141 [95%CI −0.191, 0.474], *P* = 0.402); drink occasionally compared with non-drinker (−0.001 [95%CI −0.303, 0.300], *P* = 0.993); drink daily, compared with non-drinker (−0.387 [95%CI −0.820, 0.045], *P* = 0.079); ex-smoker, compared with non-smoker (0.249 [95%CI −0.109, 0.607], *P* = 0.172); current smoker, compared with non-smoker (−0.127 [95%CI −0.816, 0.563], *P* = 0.718); having any comorbidities (0.087 [95%CI −0.311, 0.485], *P* = 0.668); and body mass index (0.009 [-0.033, 0.050], *P* = 0.674).

**Fig. 4.**
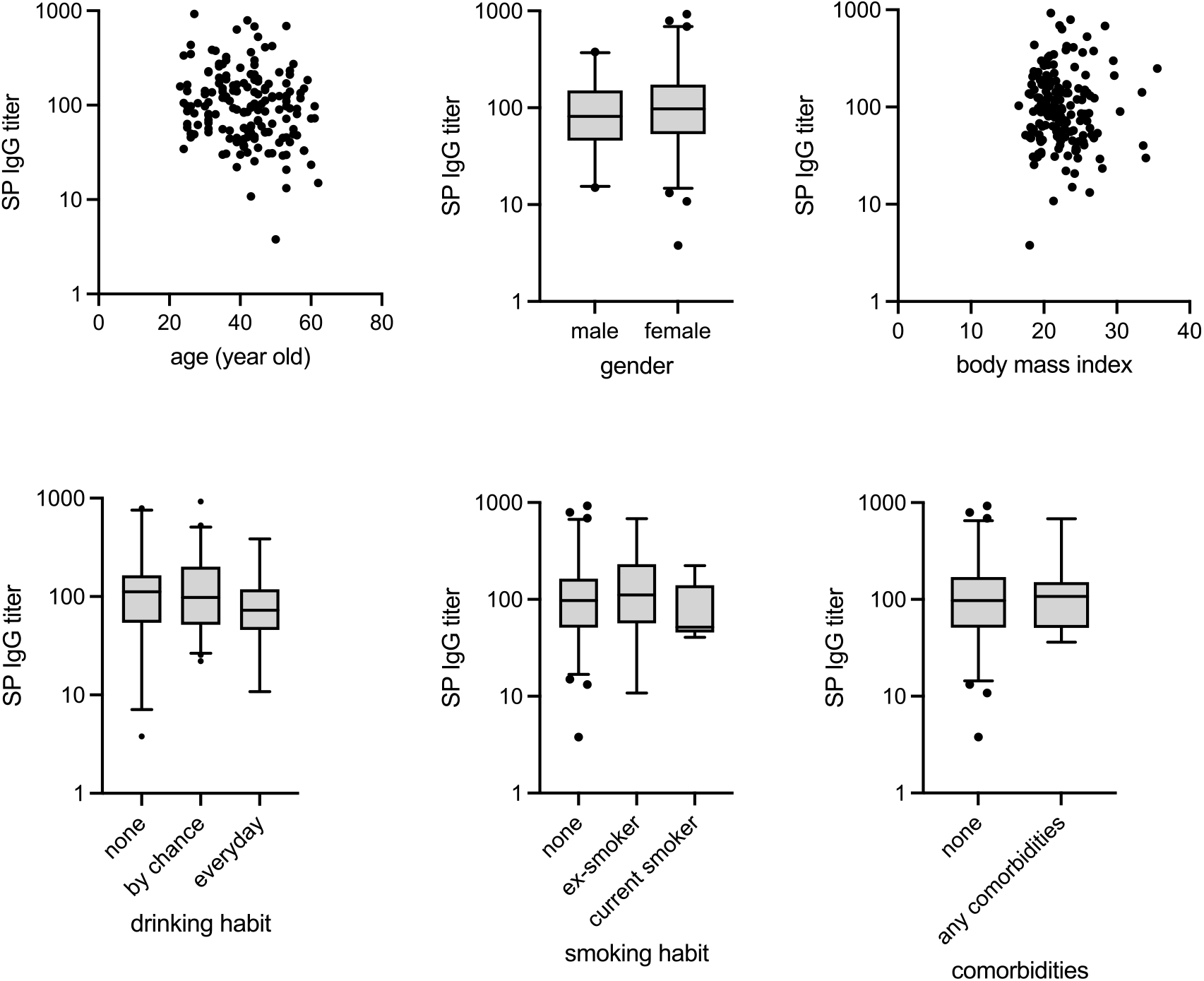
SP IgG titers at 6 weeks after the first dose of BNT162b2 and background characteristics of the 168 subjects. The bars represent median [IQR] and the error bars represent 95% confidence intervals.

## Discussion

We utilized an automated chemiluminescent enzyme immunoassay (AIA-CL reagent) to investigate the humoral immune response after vaccination against SARS-CoV-2. The AIA-CL method provided good index values of the antibody response after the BNT162b2 vaccination. This method is capable to process a large number of samples rapidly. Our data indicate that SP IgG index values in subjects who had completed two doses of the BNT162b2 vaccine had moderate correlation with neutralizing activities using lentivirus-based pseudovirus assays. This method can be adopted to evaluate subjects that have received different types of COVID-19 vaccines; it can also be used to screen for herd immunity. The BNT162b2 vaccine seemed to induce sufficient immunity at 3 weeks after the second vaccination, not 1 week after the second dose. Our results show that individuals who received only one dose of BNT162b2 did not have sufficient humoral immunity. Currently, the possibility of breakthrough infections after vaccination is raising concerns. According to a case series report, the neutralizing antibody titers in people who developed breakthrough infection were significantly lower than those without breakthrough infection (177.7 vs 501.3) [12]. In this study, a neutralizing antibody index of 100 or an AIA-CL SP IgG index of 10 were observed in the fully vaccinated individuals. However, these indexes should be carefully interpreted and should not be considered as a protective antibody titer. Higher values of SP IgG index and neutralizing antibody titers are required to lower the risk of developing COVID-19 [11].

We also detected a difference in the humoral immune responses elicited by BNT162b2 against the original strain and the variants of concern using the prototype reagent panels. Our results demonstrate that the levels of the SP IgG index against the Beta variant were 54% of those of the original strain. By contrast, the SP IgG index value for the Alpha strain was double that of the original strain. Another study reported that the antibody titer against Alpha strain induced by vaccination was not different with that against the original strain [13]. We suggest that the chemiluminescent enzyme immunoassay can capture the difference in humoral responses to the variant strains such as the Beta strain, and this approach can be adopted as a mass screening method.

Several background factors affected the SP IgG index titers in fully vaccinated subjects. We found that older age may be related to a lower antibody response. Regarding age, vaccinated individuals over 80 years old tend to have lower antibody titers [14,15]. Another report looking at a Japanese population concluded that the antibody titers in the subjects who drink alcohol frequently are lower than those of other cohorts in Japan [16]. The effect of BMI on the antibody titer after vaccination is still controversial. The antibody titer against the SP acquired by natural infection is higher in severely obese patients (BMI > 25) [17] compared to those with BMI <18.5. By contrast, another report suggests that obesity is associated with low antibody titer after vaccination [18,19]. In our study, drinking habit and BMI did not affect SP IgG index titers. Further research is required to investigate the relationship between individual characteristics and antibody titers induced by BNT162b2 vaccination. Our results suggest that humoral immunity after vaccination varies with the individual. Regardless of individual characteristics, we strongly suggest completing two doses of vaccination for people who have had no history of COVID-19 infection.

Our study had several limitations. First, our study cohort, consisting of 168 subjects, is small. Cohort size can affect the relationship between predisposing factors and antibody titers. Second, we did not evaluate cell-mediated immunity in our study. SARS-CoV-2 vaccines induce both cell-mediated and humoral immunity [20]. Thus, evaluating cell-mediated immunity contributes to a more thorough analysis of vaccine effectiveness.

## Data Availability

The data that support the findings of this study are available from the corresponding author upon reasonable request.

## Conflict of Interest

NO is an employee of Tosoh Corporation. YY is an employee of Kanto Chemical Co., Inc. Other authors stated no conflict of interest.

## Author contributions

HK contributed to the study design, data collection, statistical analysis and interpretation of data, and the drafting and editing of the manuscript. HG, SY, and TS contributed to the study design and data collection. KM, NO, and YY performed the laboratory tests. TM, AG, HN, and AR contributed to the study design, data collection, and supervised the analysis and preparation of the manuscript. All authors made critical revisions to the manuscript for important intellectual content and approved the final manuscript. All authors meet the ICMJE authorship criteria.

## Acknowledgments

The authors thank all the participants and medical staff for their participation and assistance in the study, and Dr. Yuichiro Yano and Dr. Yusuke Kobayashi at YCU Center for Novel and Exploratory Clinical Trials for their valuable advice, and Misa Katayama at Yokohama City University for secretarial assistance. This work was supported by the Japan Agency for Medical Research and Development (AMED) under Grant Number: JP20he0522001. The authors would like to thank Enago (www.enago.jp) for the English language review.

## Notes

### Author Declarations

This study was approved by the institutional review board of Yokohama City University Hospital (Approval number: B210300001).

